# Beneficial effects of colchicine for moderate to severe COVID-19: an interim analysis of a randomized, double-blinded, placebo controlled clinical trial

**DOI:** 10.1101/2020.08.06.20169573

**Authors:** Maria IF Lopes, Letícia P Bonjorno, Marcela C Giannini, Natália B Amaral, Maíra N Benatti, Uebe C Rezek, Laerte L Emrich Filho, Betânia AA Sousa, Sérgio CL Almeida, Rodrigo Luppino-Assad, Flávio P Veras, Ayda Schneider, Tamara S Rodrigues, Luiz OS Leiria, Larissa D Cunha, José C Alves-Filho, Thiago M Cunha, Eurico Arruda, Carlos H Miranda, Antonio Pazin-Filho, Maria A Martins, Marcos C Borges, Benedito AL Fonseca, Valdes R Bollela, Cristina M Del-Ben, Fernando Q Cunha, Dario S Zamboni, Rodrigo C Santana, Fernando C Vilar, Paulo Louzada-Junior, Renê DR Oliveira

## Abstract

**Introduction:** Neutrophilia and high levels of proinflammatory cytokines and other mediators of inflammation are common finds in patients with severe acute respiratory syndrome due to COVID-19, a dramatic condition for which there is no specific treatment, but supportive care and attempts to control the systemic inflammation. By its action on leukocytes, we propose colchicine as an intervention worthy of being tested.

**Objective:** To evaluate whether the addition of colchicine to standard treatment for COVID-19 results in better outcomes.

**Methods:** We present the interim analysis of a single-center randomized, double-blinded, placebo controlled clinical trial of colchicine for the treatment of moderate to severe COVID-19, with 38 patients allocated 1:1 from April 11 to July 06, 2020. Colchicine regimen was 0.5 mg thrice daily for 5 days, then 0.5 mg twice daily for 5 days. The first dose was 1.0 mg whether body weight was ≥ 80 kg.

**Endpoints:** The primary endpoints were the need for supplemental oxygen; time of hospitalization; need for admission and length of stay in intensive care unit; and death rate and causes of mortality. As secondary endpoints, we assessed: serum C-reactive protein, serum Lactate dehydrogenase and relation neutrophil to lymphocyte of peripheral blood samples from day zero to day 7; the number, type, and severity of adverse events; frequency of interruption of the study protocol due to adverse events; and frequency of QT interval above 450 ms.

**Results:** Thirty-five patients (18 for Placebo and 17 for Colchicine) completed the study. Both groups were comparable in terms of demographic, clinical and laboratory data at baseline. Median (and interquartile range) time of need for supplemental oxygen was 3.0 (1.5-6.5) days for the Colchicine group and 7.0 (3.0-8.5) days for Placebo group (p = 0.02). Median (IQR) time of hospitalization was 6.0 (4.0-8.5) days for the Colchicine group and 8.5 (5.5-11.0) days for Placebo group (p = 0.03). At day 2, 53% vs 83% of patients maintained the need for supplemental oxygen, while at day 7 the values were 6% vs 39%, in the Colchicine and Placebo groups, respectively (log rank; p = 0.01). Hospitalization was maintained for 53% vs 78% of patients at day 5 and 6% vs 17% at day 10, for the Colchicine and Placebo groups, respectively (log rank; p = 0.01). One patient per group needed admission to ICU. No recruited patient died. At day 4, patients of Colchicine group presented significant reduction of serum C-reactive protein compared to baseline (p < 0.001). The majority of adverse events were mild and did not lead to patient withdrawal. Diarrhea was more frequent in the Colchicine group (p = 0.17). Cardiac adverse events were absent.

**Discussion:** The use of colchicine reduced the length of both, supplemental oxygen therapy and hospitalization. Shortly less than half of the patients of the Colchicine group stopped receiving supplemental oxygen until day 2. Clinical improvement was in parallel with a reduction on serum levels of C-reactive protein. The drug was safe and well tolerated. Colchicine may be considered a beneficial and not expensive option for COVID-19 treatment. Clinical trials with larger numbers of patients should be conducted to further evaluate the efficacy and safety of colchicine as an adjunctive therapy for hospitalized patients with moderate to severe COVID-19.

## INTRODUCTION

Systemic inflammation is the hallmark of moderate to severe cases of coronavirus-19 disease (COVID-19) [1]. Its outbreak has already sent millions to infirmaries and intensive care units (ICU) throughout the world, mainly due to pulmonary infiltrates resulting in the severe acute respiratory syndrome (SARS) [2]. High levels of interleukin (IL)-1β, IL-6, IL-18 and tumor necrosis factor (TNF) are some of the many immunologic disturbances in the pathophysiology of the high inflammatory status of COVID-19 [3], which counts, moreover, with markedly elevation of serum C-reactive protein (CRP) and uncommon neutrophilia and lymphopenia [4, 5]. Neutrophils release neutrophil extracellular traps (NETs), which were found to be toxic to lung epithelial cells *in vitro*. Furthermore, high levels of NETs were present in the plasma of COVID-19 patients compared to healthy controls and the presence of these cellular components was at least ten times higher in tracheal aspirates than in plasma of the same patients, raising the question whether they have a role in the lung lesions [6 and under revision elsewhere].

The inflammasome of NOD-, LRR- and pyrin domain-containing protein 3 (NLRP3) may be important in certain antiviral responses [7]. After viral activation of the protein complex, mainly in monocytes and antigen-presenting tissue cells, its constituent pro-caspase-1 suffers auto-cleavage and, by its turn, cleaves pro-IL-1β and pro-IL-18 to their active form – IL-1β and IL-18 [8]. Both products activate B, T and NK cells in addition to stimulating the release of other inflammatory cytokines [9]. It seems appropriate to infer that an aberrant activation of inflammasome underlies the “hyper” inflammation found in hospitalized COVID-19 patients.

For decades, colchicine has been successfully used for the treatment and prevention of crystal-induced arthritis, e.g. gout. Systemic auto-inflammatory diseases such as familial Mediterranean fever and Behçet’s disease are conditions in which colchicine use may be necessary continuously [10]. Much of this success comes from its direct effect on phagocytes residing or migrating into the synovial joints, vessel walls or other tissues, leading to inflammasome inhibition and impaired production and release of IL-1β [11] and NETs [12]. In all of these situations, the drug is well tolerated and its adverse effects are broadly recognized.

Piantoni *et al* [13] discussed the rationale of its use for the treatment of COVID-19, with focus on the control of the systemic inflammation caused by SARS-CoV-2 infection. A case-control study [14] and an open-label clinical trial [15] of colchicine for COVID-19 have results published hitherto.

## METHODS

### Trial design

We conducted a randomized, double-blinded, placebo controlled clinical trial to evaluate the use of colchicine for the treatment of hospitalized patients with moderate to severe COVID-19. The randomization was performed 1:1 for placebo or colchicine by using the online tool at https://www.randomizer.org/.

Taking into account the prominent need of efficient therapies for COVID-19, besides limitations to conduct a clinical trial in a single center, 60 patients seemed to be appropriate whether randomized into two groups of 30 patients.

The trial is registered on the National Registry under the alphanumeric code RBR-8jyhxh (http://www.ensaiosclinicos.gov.br/rg/RBR-8jyhxh/) and it was approved by National Ethics Committee (CONEP; CAAE: 30248420.9.3001.5403). All patients signed the Consent Form.

### Intervention

Patients of the intervention arm received colchicine 0.5 mg thrice daily for 5 days, then 0.5 mg twice daily for 5 days; if body weight ≥ 80 kg, the first dose was 1.0 mg. Whether a patient had chronic kidney disease, with glomerular filtration rate under 30 mL/min/1.73 m^2^, colchicine dose was reduced to 0.25 mg thrice daily for 5 days, then 0.25 mg twice daily for 5 days, no matter the body weight.

All participants received the institutional treatment for COVID-19 with azithromycin 500 mg once daily for up to 7 days, hydroxychloroquine 400 mg twice daily for 2 days, then 400 mg once daily for up to 8 days and unfractionated heparin 5000 UI thrice daily until the end of hospitalization. Methylprednisolone 0.5 mg/kg/day for 5 days could be added if the need for supplemental oxygen was 6 L/min or more. Study and institutional protocol drugs were suspended when participants reached good clinical and laboratory parameters and could be discharged. Study medication was suspended when a patient needed ICU admission, following local restrictions for its use in critical patients.

### Study population

The inclusion criteria were: individuals hospitalized with moderate or severe forms of COVID-19 [16] diagnosed by RT-PCR in nasopharyngeal swab specimens and lung computed tomography scan involvement compatible with COVID-19 pneumonia; older than 18 years; body weight > 50 kg; normal levels of serum Ca^2+^ and K^+^; QT interval < 450 ms at 12 derivations electrocardiogram (according to the Bazett formula) and negative serum or urinary β-HCG if woman under 50. The exclusion criteria were defined as: mild form of COVID-19 or in need for ICU admission; diarrhea resulting in dehydration; known allergy to colchicine; diagnosis of porphyria, myasthenia gravis or uncontrolled arrhythmia at enrollment; pregnancy or lactation; metastatic cancer or immunosuppressive chemotherapy; regular use of digoxin, amiodarone, verapamil or protease inhibitors; chronic liver disease with hepatic failure; inability to understand the Consent Form.

### Setting

Patients were evaluated daily and blood collection for general laboratory tests were performed at days zero, 2, 4 and 7 if discharge did not happen before. Twelve derivations ECG were performed each 24-48 hours.

### Main endpoints

The primary endpoints were clinical parameters, such as the time of need for supplemental oxygen; time of hospitalization; need for admission and length of stay in ICU; and death rate and causes of mortality. As secondary endpoints we assessed clinical and laboratory parameters: measures of serum CRP, serum LDH and relation neutrophil to lymphocyte of peripheral blood samples from day zero to day 7; the number, type, and severity of adverse events; frequency of interruption of the study protocol due to adverse events; and frequency of QT interval above 450 ms.

### Statistical analysis

We present descriptive statistics as absolute numbers and percentage or median and interquartile range (IQR). Absolute numbers and percentage were compared with Fisher’s exact test. Comparisons of clinical and laboratory parameters expressed in median and IQR were done through Mann-Whitney test. Additionally, Kaplan-Meier survival curves were performed, with analysis by Mantel-Haenszel log rank test, to compare the time to abandon supplemental oxygen and time to discharge between the groups. Kruskall-Wallis test was used for comparisons of laboratory exams at the four blood collection times, followed by Dunn’s Multiple Comparison test. For all tests, p < 0.05 was considered for statistical significance.

## RESULTS OF INTERIM ANALYSIS

### Participants

The enrollment started on April 11 and stopped on July 6, 2020. We assessed 72 patients and included 38 for randomization as shown in **Figure 1**. We programmed a first interim analysis after the study completion for the 30th randomized patient. For that occasion, we found significant differences between the groups for the primary endpoints. Then, we resumed the enrollment up to 38 patients and for this new interim analysis the difference resulted sustained.

**Figure 1.**
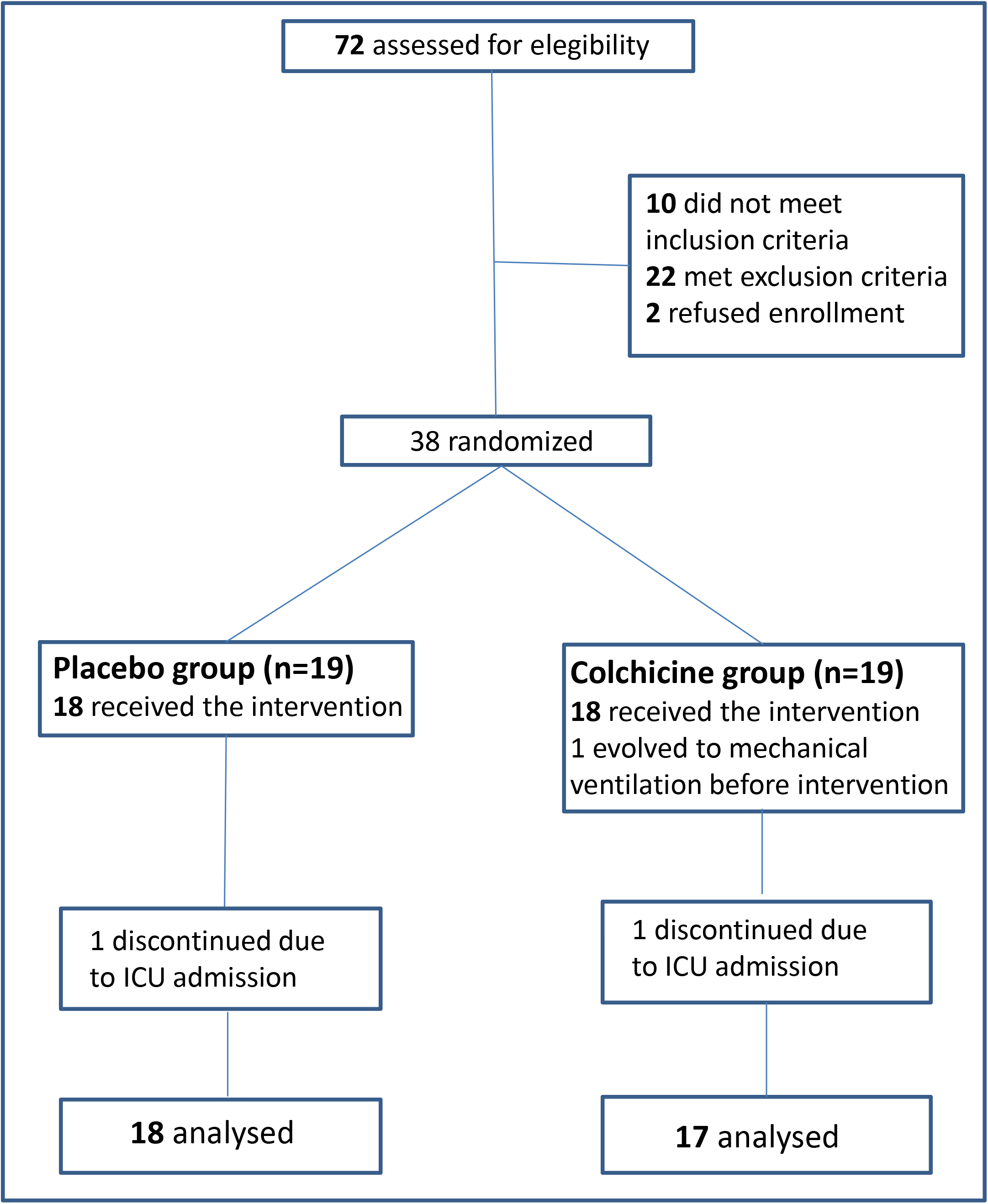
Study Flow Diagram

The baseline laboratory and clinical characteristics of the 35 patients who completed the study are presented in **Table 1**. All patients received the institutional protocol treatment with hydroxychloroquine, azithromycin and heparin. Seven patients in each group received methylprednisolone. No treatment, institutional or interventional, was interrupted due to adverse events. The groups were similar in terms of demographic characteristics, clinical status and laboratory evaluation at baseline. Fatigue was more common and the creatinine level was numerically greater in the Colchicine group, although no clinical significance should be awarded for them. There was a slight predominance of men in the Colchicine group. Moreover, all patients in the Colchicine group had body mass index (BMI) above 25.0 kg/m^2^ (data not shown) and the group median PaO_2_/FiO_2_ was 55.0 lower, for both parameters with no statistical difference.

**Table 1.** COVID-19 patients characteristics at baseline

|  | Placebo group (n=18) | Colchicine group (n=17) | p-value |
| --- | --- | --- | --- |
| **Demographics** |  |  |  |
| Men [n (%)] | 5 (27.8) | 9 (52.9) | 0.17 |
| Age [years; median (IQR)] | 53.5 (35.5-65.5) | 48.0 (41.5-64.0) | 0.94 |
| Time of symptoms [days; median (IQR)] | 7 (6.5-9.0) | 9 (7.0-10.5) | 0.08 |
| **Comorbidities** |  |  |  |
| Smoking currently or formerly [n (%)] | 5 (28) | 2 (12) | 0.40 |
| Respiratory diseases [n (%)] | 2 (11) | 3(18) | 0.66 |
| Cardiovascular diseases [n (%)] | 6 (33) | 8 (47) | 0.50 |
| *Diabetes mellitus* [n (%)] | 6 (33) | 5 (29) | 1 |
| Dyslipidemia [n (%)] | 6 (33) | 3 (18) | 0.44 |
| BMI [kg/m<sup>2</sup>; median (IQR)] | 30.6 (26.8-34.1) | 33.9 (30.0-39.9) | 0.21 |
| **Clinical Picture of COVID-19** |  |  |  |
| Fever [n (%)] | 18 (100) | 17 (100) | 1 |
| Cough [n (%)] | 18 (100) | 17 (100) | 1 |
| Fatigue [n (%)] | 5 (28) | 12 (70) | **0.01** |
| Myalgia [n (%)] | 9 (50) | 8 (47) | 1 |
| Diarrhea [n (%)] | 5 (28) | 7 (41) | 0.49 |
| PaO<sub>2</sub>/FiO<sub>2</sub> at enrollment [median (IQR)] | 284 (216-339) | 229 (176-307) | 0.12 |
| qSOFA ≥1 [n (%)] | 16 (89) | 16 (94) | 1 |
| SOFA | 2.5 (1-3) | 2.0 (1-3) | 0.85 |
| **Laboratory findings** |  |  |  |
| Hemoglobin [g/dL; median (IQR)] | 12.6 (11.3 – 14.2) | 13.2 (12.2 – 14.8) | 0.20 |
| Neutrophils [/mm<sup>3</sup>; median (IQR)] | 4650 (3300-7050) | 4800 (3050-6200) | 0.81 |
| Lymphocytes [/mm<sup>3</sup>; median (IQR)] | 1250 (680-1850) | 1400 (900-1950) | 0.40 |
| Neutrophil/Lymphocyte [median (IQR)] | 3.38 (2.25-6.25) | 2.83 (2.43-4.39) | 0.39 |
| Platelets [/mm<sup>3</sup>; median (IQR)] | 247000 (170000-285000) | 218000 (201000-356000) | 0.71 |
| Creatinine [mg/dL; median (IQR)] | 0.70 (0.57-0.81) | 0.90 (0.65-0.99) | **0.03** |
| C-reactive protein [mg/dL; median (IQR)] | 8.2 (5.5-14.3) | 7.8 (5.6-10.3) | 0.56 |
| Lactate dehydrogenase [U/L; median (IQR)] | 317 (262-433) | 339 (274-474) | 0.68 |
| D-dimer [mcg/mL; median (IQR)] | 0.85 (0.58-1.64) | 1.02 (0.85-1.53) | 0.28 |
| Ferritin [ng/mL; median (IQR)] | 565 (215-1126) | 711,7 (436-1266) | 0.18 |
| Aspartate aminotransferase [U/L; median (IQR)] | 30 (25-53) | 47 (29-76) | 0.19 |
| Alanine aminotransferase [U/L; median (IQR)] | 36 (26-61) | 50 (34-114) | 0.20 |
| **Medications** |  |  |  |
| Hydroxychloroquine [n (%)] | 18 (100) | 17 (100) | 1 |
| Azithromycin [n (%)] | 18 (100) | 17 (100) | 1 |
| Unfractionated heparin [n (%)] | 18 (100) | 17 (100) | 1 |
| Methylprednisolone [n (%)] | 7 (39) | 7 (41) | 1 |

*IQR – Interquartile range; BMI – Body mass index; PaO<sub>2</sub> – Arterial oxygen partial pressure; FiO<sub>2</sub> – Fractional inspired oxygen; SOFA – Sequential Organ Failure Assessment Score; qSOFA – quick SOFA; D-dimer – Dimerized plasma fragment D*

### Outcomes

One patient per group needed admission to ICU. Although the interventions were stopped, both patients were followed for outcomes: the time of treatment before ICU admission was 2 and 3 days, the length of stay in ICU was 12 and 11 days, and the time of hospitalization was 23 and 26 days, respectively, for Colchicine and Placebo groups. No recruited patient died.

The need for supplemental oxygen and the time of hospitalization are shown in **Table 2**. The median for both parameters was lower for the Colchicine group compared to the Placebo group. Half of the patients receiving colchicine stopped using supplemental oxygen for the third day of intervention, while the same happened to half of the patients receiving placebo on the seventh day (p = 0.02). Significant difference (p = 0.03) between the groups was found for time of hospitalization, in detriment of the Placebo group.

**Table 2.** Primary Outcomes

| Outcome | Placebo group (n=18) | Colchicine group (n=17) | p-value |
| --- | --- | --- | --- |
| Time of supplemental O<sub>2</sub> [days; median (IQR)] | 7.0 (3.0-8.5) | 3.0 (1.5-6.5) | **0.02** |
| Time of hospitalization [days; median (IQR)] | 8.5 (5.5-11.0) | 6.0 (4.0-8.5) | **0.03** |

*IQR – Interquartile range*

Kaplan-Meier survival curves for the need for supplemental oxygen and the maintenance of hospitalization are depicted in **Figure 2** and **Supplementary Figure 1**, respectively. At day 2, 53% vs 83% of patients maintained the need for supplemental oxygen, while at day 6 these values were 24% vs 56%, in the Colchicine and Placebo groups, respectively (log rank test, 6.25; p = 0.01). Hospitalization was maintained for 41% vs 72% of patients at day 6 and 6% vs 28% at day 10, in the Colchicine and Placebo groups, respectively (log rank test, 5.55; p = 0.01). Both outcomes presented similar behavior, once the last is at large extension a consequence of the first one.

**Figure 2.**
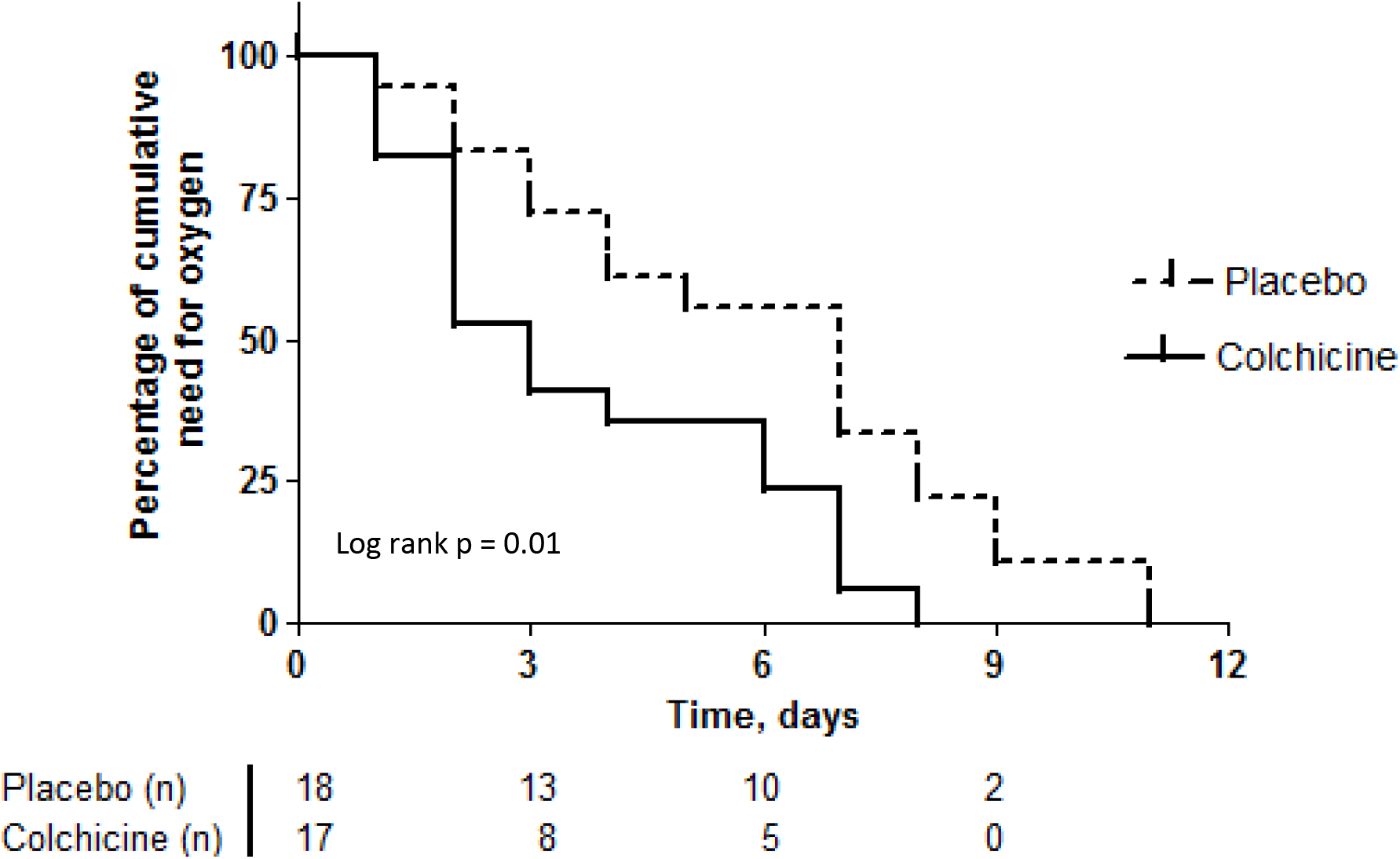
Kaplan-Meier curves of time to the end of need for supplemental oxygen for both groups

We evaluated some laboratory parameters as secondary endpoints. **Figure 3** and **Supplementary Figures 2 and 3** show the temporal variations of serum CRP and LDH, and peripheral blood relation neutrophil to lymphocyte, respectively, from day zero to day 7. Starting both groups at similar levels of serum CRP, at day 4 patients of the Colchicine group presented significant reduction compared with themselves and with patients of the Placebo group at day zero (p < 0.001). It is possible to observe that serum levels of CRP became different between the groups in the interval between days 2 and 4, with return to normal range (median < 0.5 mg/dL) for the Colchicine group at day 4. For the Placebo group, statistical difference compared to the baseline occurred at day 7 (p < 0.01), but no return to normal range of median CRP was observed at that time. The post-test for LDH showed a difference between day zero and day 4 for the Colchicine group. No difference inter and intragroup for the relation neutrophil to lymphocyte was obtained.

**Figure 3.**
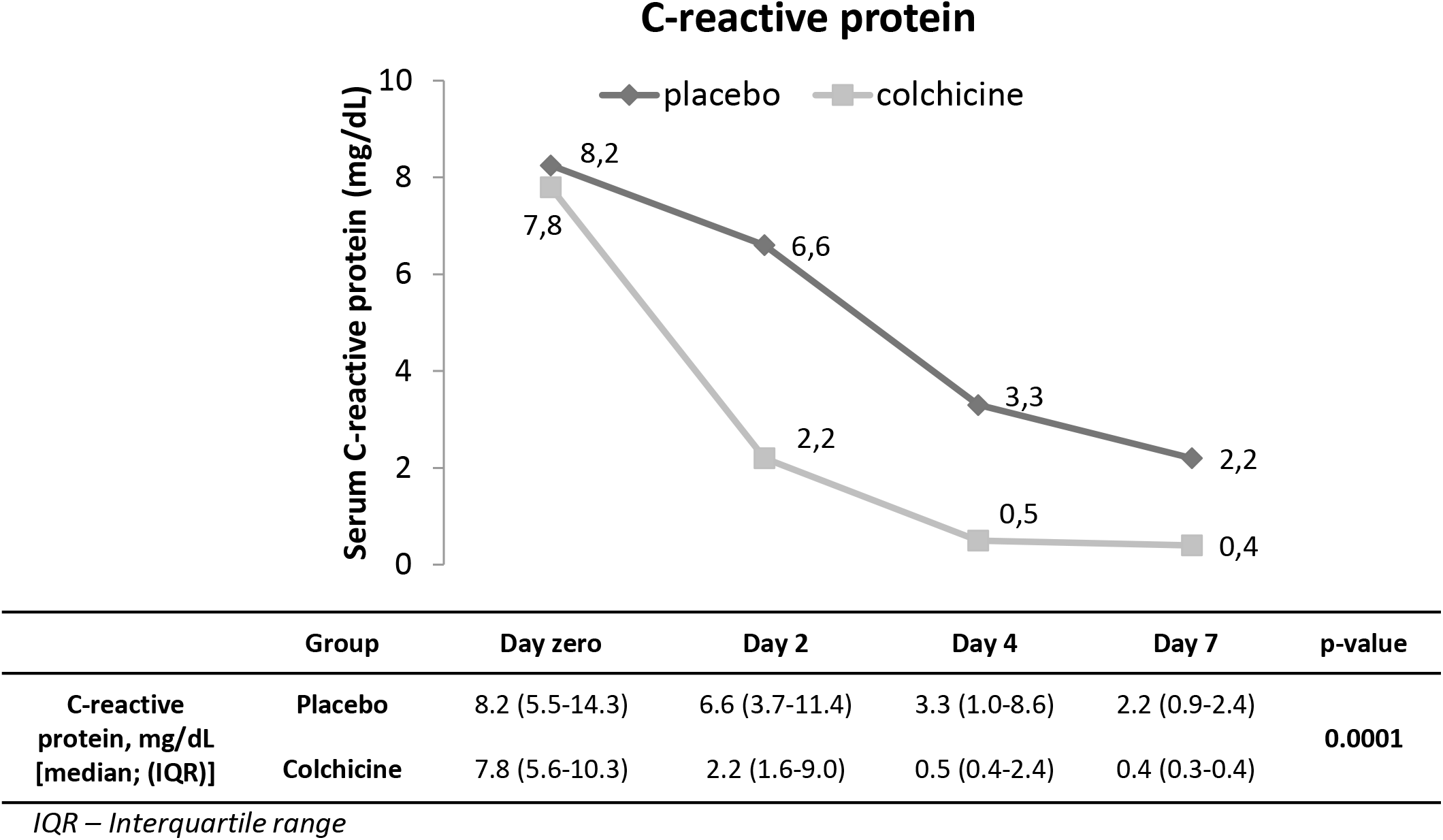
Temporal variation of serum C-reactive protein from D zero to D7 for both groups

The majority of adverse events (**Table 3**) was mild (exception for pneumonia) and, to some extent, attributable to the viral infection itself or its complications, not entailing patients withdrawal. It seems to be the case of AST and ALT transient elevations, under 3X the upper limit of normal, with no difference between the groups (data not shown). New or worsened diarrhea was more frequent in the intervention group (24% vs 6%). None of the patients suffered dehydration and the diarrhea was controlled with the prescription of an antisecretory agent (e.g., racecadotril). Cardiac adverse events, undoubtedly the main issue on the use of hydroxychloroquine and/or azithromycin for COVID-19, did not have an augmented frequency by adding colchicine. No participant had QT interval above 450 ms during the observational period (data not shown). The difference between the groups on QT interval variation (p = 0.05) from the value of day zero to the highest value does not seem to have a clinical significance, once no patient presented any cardiovascular signs or symptoms.

**Table 3.** Adverse events

| **Adverse event** | **Placebo group (n=18)** | **Colchicine group (n=17)** | **p-value** |
| --- | --- | --- | --- |
| AST transient elevation [n (%)] | 1 (6) | 1 (6) | 1 |
| ALT transient elevation [n (%)] | 3 (17) | 4 (24) | 1 |
| Nausea/vomiting [n (%)] | 3 (17) | 1 (6) | 0.6 |
| Abdominal pain [n (%)] | 2 (11) | 2 (12) | 1 |
| New or worsened diarrhea [n (%)] | 1 (6) | 4 (24) | 0.17 |
| Nosocomial pneumonia [n (%)] | 2 (11) | 2 (12) | 1 |
| Arrhythmia [n (%)] | 0 | 0 | 1 |
| QT interval at day 3 [ms; median (IQR)] | 401 (390-407) | 407 (395-418) | 1 |
| QT interval variation [ms; median (IQR)] | 33 (13-40) | 20 (9-26) | 0.05 |
| Death | 0 | 0 | 1 |

*AST – aspartate aminotransferase; ALT – alanine aminotransferase; IQR – Interquartile range*

## DISCUSSION

We presented the partial results of a randomized, double-blinded, placebo controlled clinical trial of colchicine for COVID-19. Patients receiving colchicine abandoned oxygen supplementation earlier than those receiving placebo (median, 3.0 vs 7.0 days) and the time until discharge followed a similar tendency. These two primary endpoints have relevance for daily practice in the COVID-19 pandemic, by reducing the length of hospitalization, consequently diminishing costs and the need for hospital beds. Besides that, the treatment with colchicine is not expensive.

Mainly women obese represented the study population. A wide variation of the magnitude of systemic inflammation, reflected in the broad ranges of serum CRP (2.3 to 24.4 mg/dL) and SatO_2_/FiO_2_ (77 to 400), assures that this sample of patients is widely representative of the hospitalizations due to COVID-19 pneumonia. Taking into account the effect on both, the CRP curve and the need for supplemental oxygen, colchicine use seems promising if we consider that the systemic inflammation was safely halted in a shorter period compared to the standard treatment.

The two patients who needed ICU admission had SatO_2_/FiO_2_ under 100 at enrollment. It is certain that the effect of colchicine in reducing systemic inflammation is not enough for every patient and that for some of them no intervention would prevent respiratory failure. These two patients received methylprednisolone. The study protocol used the maximum safe daily dose of colchicine [17] considering a body weight of 50 kg, i.e., 0.030 mg/kg. Body weight of the participants ranged from 62 to 145 kg, which results, for some patients, in a daily dose under 0.015 mg/kg, the minimal dose for chronic use of colchicine. This range was maintained, even for the first 24 hours, aiming to avoid the occurrence or worsening of diarrhea, a frequent manifestation of COVID-19.

Only 3 out of 35 analyzed patients had normal values of BMI, all randomized to the Placebo group. Systemic inflammation is a common characteristic linking obesity, metabolic syndrome and elevated cardiovascular risk [18, 19], with highlighted role for IL-1β and NLRP3 inflammasome. The adipose tissue of obese has macrophage type 1 as the main resident phagocytic cell. This subgroup of macrophages is primed to secrete proinflammatory cytokines, such as IL-1β, IL-6 and TNF [20]. Some groups found obesity a risk factor for severity of COVID-19, as reported in a systematic review with meta-analysis [21].

Colchicine was already tested as protective against ischemic events post myocardial infarction with some success [22] and it was found to ameliorate endothelial function in a subgroup of patients with coronary disease and augmented peripheral blood leukocyte count [23].

Recently, our group reported the possible role for neutrophil extracellular traps on lung inflammation in COVID-19 [6]. One of the actions of colchicine is to reduce migration of leukocytes, mainly neutrophils, to inflamed tissues [9]. Whatever the mechanism of action – inhibiting inflammasome, reducing neutrophil migration and activation or preventing endothelial damage –, colchicine seems to be beneficial for the treatment of hospitalized COVID-19 patients. The markedly reduction of serum CRP levels between second and fourth days coincides with clinical recovery of the majority of patients. Most of the participants were in the second week of symptoms, a phase in which systemic inflammation becomes striking. It is very unlikely that colchicine has some antiviral effect.

Worthy of note is the fact that the drug did not contribute to hepatic or cardiac adverse events nor caused immunosuppression. Diarrhea was not a limiting adverse event and an antisecretory drug may be added when necessary.

The so-called GRECCO-19 trial, an open-label study, had some results recently published [15]. The authors found that patients receiving colchicine were less prone to clinical deterioration, despite the fact that their serum levels of CRP showed no significant difference compared to those of patients not receiving the drug. For a matter of comparison, when analyzing patients with body weight > 60 kg in use of azithromycin (approximately 92%), patients of GRECCO study received a total dose of 5.0 mg of colchicine in the first 5 days, while in our study patients received 7.5 or 8.0 mg in the same period. This difference of dosage ≥ 50% for the first 5 days may explain the evident reduction of serum CRP in the present study and justify the better evolution of the intervention group. Scarsi *et al* [14] showed an association between treatment with colchicine and improved survival in a single-center cohort of adult hospitalized patients with COVID-19 pneumonia and SARS. They conclude that their results may support the rationale of colchicine use for the treatment of COVID-19 and that the efficacy and safety must be determined in controlled clinical trials. Authors used the dose of 1.0 mg/day for all patients with reduction to 0.5 mg if diarrhea occurred.

Our clinical trial has some limitations. The most evident is the reduced number of patients and the recruitment in a single center. To balance this, the blinding and the control by a placebo arm strengthen our finds. The absence of mechanistic investigations, e.g., measures of the plasmatic levels of cytokines, is another limitation. Our exclusion criteria were some kind of restrictive, such as the prohibition of some cardiovascular drugs. Much of this concern was related to drugs that could impair colchicine metabolism or excretion, but some concern was also due to the potential hazardous effect of hydroxychloroquine and azithromycin combined use for myocardial fibers. Finally, as patients were discharged, the number of blood samples reduced through the first week of observation and beyond, once it would not be appropriate to summon up the patients for new blood collections due to SARS-CoV-2 transmission possibility.

## CONCLUSIONS

Patients who received colchicine in this randomized, double-blinded, placebo controlled clinical trial presented better evolution in terms of the need for supplemental oxygen and the length of hospitalization. Serum CRP was a laboratory marker of clinical improvement. Colchicine was safe and well tolerated. Clinical trials with larger numbers of patients should be conducted to further evaluate the efficacy and safety of colchicine as an adjunctive therapy for hospitalized patients with moderate to severe COVID-19.

## Data Availability

I declare that all clinical and laboratory data presented in the manuscript are promptly available.

## Author Contributions

RDRO, RCS, FCV and PLJ contributed to the study design. RDRO contributed to randomization and administrative support. FCV, RCS, BAAS, MIFL, LPB, MCG, NBA, MNB, LLEF, SCLA, RLA, MAM, MCB, BAL and CHMJ contributed to patients’ selection and follow-up. UEC analyzed all 12 derivations ECG. MIFL, LPB, MCG, NBA, MNB, FPV, AS and TSR contributed to blood collection and processing. RDRO, MIFL, LPB, MCG, NBA. PLJ, SCLA and RLA contributed to clinical data collection. RDRO, PLJ and CMDB contributed to statistical analysis. RDRO wrote the manuscript draft. PLJ, VRB, BAF, FQC, DSZ, TMC, JCAF, LOSL, LDC, EA and CMDB revised the manuscript. FQC, JCAF, TMC, LOSL, LDC, DSZ and PLJ obtained funding. All authors approved the manuscript.

## Acknowledgments

We are grateful to Muriel C. R. O. Berti, Basílica Botelho Muniz and Livia Maria C. S. Ambrósio for technical assistance.

